# Protocol for a Multi-Methods Explanatory Evaluation of the “Healthy Homes and Neighbourhoods” Program: Impact, Implementation, and Value-for-Money in an Integrated Care Initiative

**DOI:** 10.64898/2026.11.19.26368549

**Authors:** K. Yakubu, F. Mukumbang, Ana Campain, M. Cunich, E. R. Atkins, J. Coombes, K. Hunter, K. Bennet-Brooke, T. Madden, M. McDonell, A. Thiraviarajah, J. Lin, I. Tan, S. Forell, G. Schierhout, S. Jan, C. Huckel-Schneider, P. Fotheringham, R. Lyons, B. Goodger, E. Miller, L. Haynes, S. Woolfenden, J. Eastwood, H. Liu

## Abstract

**Objectives:** To describe a multi-methods explanatory evaluation protocol for Healthy Homes and Neighbourhoods (HHAN), an integrated care initiative for families experiencing health and social disadvantage in a Sydney health district. The evaluation will examine how and in what contexts HHAN influences health and social outcomes and will assess its value for money.

**Design:** Multi-methods explanatory evaluation using linked administrative data, realist-informed interviews, yarning, and economic evaluation.

**Setting:** A Sydney health district, including place-based hubs and partner sites involved in HHAN delivery or support.

**Participants:** For the quantitative analyses, study participants will include participants of the HHAN program, linked family members in the Human Services Dataset, and a constructed counterfactual population. For the qualitative interviews and yarning, study participants will include service providers, program managers, policymakers, clients and carers involved in the HHAN program, including Aboriginal and Torres Strait Islander participants where culturally appropriate and approved.

**Methods and analysis:** Impact on health and social outcomes will be examined using linked administrative data with matching, weighting and regression-based approaches. A critical realist analysis of interview and yarning transcripts will explore program mechanisms of change and implementation conditions. Economic evaluation will estimate return on investment and cost-effectiveness where data permit. Findings will be integrated to explain patterns of impact and implementation across contexts.

**Strengths and limitations of this protocol:**

- This protocol describes an approach that combines linked administrative data, qualitative interviews, yarning, and economic evaluation to explain not only whether the HHAN program is associated with outcomes, but how, why, and in what contexts outcomes may occur.
- The design embeds Aboriginal and Torres Strait Islander leadership, an Aboriginal Advisory Committee and culturally grounded methods, supporting cultural safety, data sovereignty and interpretation at a knowledge interface.
- Use of a counterfactual population, weighting, regression modelling and sensitivity analyses strengthens causal interpretation within the constraints of a non-randomised real-world evaluation.
- As an observational evaluation conducted in open service systems, residual confounding, data linkage constraints and incomplete administrative data may limit the precision and generalisability of some estimates.
- Economic analyses will depend on the availability, completeness and comparability of cost and outcome data across government, health system and societal perspectives.

## 1. Introduction

Reducing entrenched health inequities is a core public health priority. Structural determinants, including political and economic forces, shape the uneven distribution of health risks and protective factors across populations and places. These determinants influence the occurrence of adverse childhood events and intergenerational patterns of inequity, reinforcing the need for services that respond to complex health and social needs ^1, 2^. Integrated care models are a type of service approaches that can offer such response. They can enable coordinated support across sectors that responds to the lived realities of families and communities. Evaluation of these models must provide evidence that helps governments uphold the right to health and enables partnership approaches to health and social service delivery that transcend sectoral silos ^1, 3^.

The Healthy Homes and Neighbourhoods (HHAN) program was established in 2015 as an integrated care initiative in a Sydney health district ^1, 2^. HHAN’s goals are to increase families’ access to, and engagement with, relevant health and social services, and to support integration of services across sectors. Guided by geospatial mapping to identify communities experiencing poor health outcomes, HHAN employs care coordinators, offers flexible, community-based outreach events, and works with partners across housing, education, justice, financial and legal sectors ^1-3^. The program emphasises early access to appropriate care and coordinated support for children’s physical, mental and developmental needs, as well as the mental health and wellbeing of their caregivers ^4, 5^.

Early feasibility studies of HHAN between 2020 and 2022 reported increased access to care, improved engagement, and more positive outlooks among participating families ^6-10^. Subsequent evaluation studies identified three generative mechanisms by which HHAN influence the health and wellbeing of its clients: strengthening the relational trust between families and providers; organisational commitment to integrated care (e.g., referral pathways and shared workflows); and families’ recognition of family–health connections that prompt earlier help-seeking and sustained engagement ^6^. These mechanisms operated under certain contexts, including historical perceptions of services, availability of skilled providers, leadership understanding, and client health literacy. The program theory from this previous HHAN evaluation is the foundation for the proposed evaluation. The evaluation builds on existing knowledge and examines how impact has evolved and what it means for policy and practice.

A critical realist methodology informed the initial program theories of how HHAN works ^6^. Seeing around 40% of HHAN families identify as Aboriginal and/or Torres Strait Islander peoples, it is therefore essential that subsequent evaluation draws from both Indigenous knowledges and Western scientific traditions, an approach we term knowledge-at-the-interface ^11, 12^. This position Indigenous and Western paradigms in respectful dialogue to inform question framing, data collection and interpretation, governance, and translation. Ongoing cycles of learning and adaptation, with attention to how key concepts are communicated across the team, are central to this approach.

### 1.1 Aims and objectives

This protocol describes an evaluation designed to: (1) examine for why, how, and in what contexts HHAN has an impact on selected health and social outcomes; (2) identify which mechanisms, operating in particular contexts, enable or constrain implementation and what this implies for sustainability and scale; and (3) assess value-for-money, focusing on return on investment (ROI) from a government perspective and cost-effectiveness from a health-sector perspective, with exploratory analyses from broader societal perspectives where feasible. This manuscript reports the protocol before completion of the planned impact, implementation and economic analyses. As such, no outcome findings are reported. The protocol’s contribution is to make explicit a prospective evaluation design for a complex, place-based integrated care program before outcome results are known, linking impact, implementation, cultural governance and economic value in a single explanatory approach.

## 2. Methods

### 2.1 Study design and setting

We will undertake a multi-methods explanatory evaluation organised around three objectives: impact, implementation enablers/constraints, and value-for-money. The study is situated within a Sydney health district and partner agencies delivering HHAN. We will analyse routinely collected and linked administrative data alongside qualitative interviews and yarning. As this is a protocol manuscript, completed-study reporting checklists are not applied here; however, the design anticipates the information needed for future reporting of the observational, routinely collected data, economic evaluation and qualitative components.

### 2.2 Participants and sampling

#### Objective 1 Impact cohort (quantitative)

As this is a non-randomised evaluation, we will construct a counterfactual population from the Health Services Dataset (HSDS) ^13^, using matching, weighting, and regression-based approaches to reduce observed confounding.

#### Objectives 1 & 2

Qualitative and yarning sampling: Based on a prior study in this program ^6^, we anticipate a minimum of 15 interviews and yarning sessions across system leaders/administrators, service providers, and clients/carers. The final sample size will be guided by information power ^14^ (i.e., we will analyse data concurrently with data collection and continue sampling until additional interviews/yarns contribute little or no new information relevant to the study aims and planned analysis, considering the specificity of participants to the evaluation questions, and the quality of interactions).

#### Objective 3

Economic evaluation: No formal sample size calculation is required. Analyses will use annual HHAN program cost data as well as linked health and social services data.

### 2.3 Eligibility

#### Quantitative impact and economic evaluation (Objectives 1 & 3)

All HHAN participants and their linked family members within HSDS will be included in the quantitative evaluation of health and social outcomes and in the economic analyses, consistent with the ethics approval.

#### Qualitative interviews and yarning (Objectives 1 & 2)

Eligible participants include all HHAN-recognised service providers across health and non-health sectors; program managers; relevant policymakers; and clients and carers (including carers participating on behalf of clients with language, cognitive or health barriers). Families declining HHAN care coordination will be excluded.

### 2.4 Recruitment

Potential participants will be approached using culturally appropriate materials (flyers, participant information statements), distributed via email/post and at place-based hubs and partner sites. Non-responders may be contacted up to two times. For Aboriginal and Torres Strait Islander participants, recruitment and informed consent processes will be guided by culturally safe practice and relational principles that prioritise comfort and mutual meaning-making.

### 2.5 Patient and public involvement

Clients, carers, service providers and community partners will contribute through qualitative interviews, yarning sessions and partner engagement processes. Aboriginal and Torres Strait Islander investigators and an Aboriginal Advisory Committee will guide study priorities, interpretation of findings and dissemination involving Aboriginal and Torres Strait Islander peoples. These processes are intended to ensure that the evaluation remains relevant to families, culturally safe, and useful for service optimisation and policy decision-making.

### 2.6 Data sources and outcomes

#### Data sources

1. **Human Services Dataset (HSDS):** a linked administrative data asset that includes domains such as health (e.g., hospital admissions, emergency department (ED) presentations), justice, education (e.g., student enrolments), and community services (e.g., housing) 13. The protocol specifies that the evaluation will document linkage approvals, data access arrangements, variable definitions, coding decisions and any restrictions on reproducibility arising from secure-data governance, so these can be reported transparently when study findings are submitted.
2. **Routine HHAN program data:** including Patient-Reported Experience/Outcome Measures (PREMs/PROMs) and an inventory of unmet needs (Appendix 1 and 2).
3. **Annual program costs:** budget statements and financial records required for return-on-investment (ROI) and cost-effectiveness analysis (CEA), and published unit costs for healthcare and social care utilisation (e.g., emergency department presentations, hospital admissions, Out-of-home care placements and restorations).

#### Outcomes

- **Health outcomes (Objective 1):** hospitalisation trends for key diagnoses (children’s physical, mental, developmental; caregivers’ mental health), ED presentations, length of stay (as available).
- **Social outcomes (Objective 1):** out-of-home care placements and restorations; selected education and justice indicators subject to data availability and approvals.
- **Implementation outcomes (Objective 2):** program reach, adoption, acceptability, appropriateness/adaptability, and staff experiences related to delivery over time (from PREMs/PROMs, qualitative interviews and yarning sessions).
- **Economic outcomes (Objective 3):** ROI ratio from a government perspective (intervention costs vs hospitalisation cost offsets), and incremental cost-effectiveness ratios (ICERs, e.g., $ per admission averted) from the health-sector perspective; broader government/societal costs and outcomes explored where feasible.

### 2.7 Quantitative analysis (Objective 1; contributes to Objective 3)

We will produce descriptive statistics for the HHAN cohort and counterfactual comparators (demographics; clinical and service-use profiles). Health and social outcomes will be modelled using weighted random-effects Poisson or negative binomial models as appropriate for count data. Where weighting is insufficient, double-adjustment methods will be applied. Longitudinal plots will examine trends over time. Sensitivity analyses will assess robustness to modelling choices and specification. All analyses will occur within the HSDS secure access environment per governance and ethics requirements; software will include SAS (9.4+), Stata (18+), and R (4.5.2+) ^15-17^. The planned analyses will pre-specify the cohort definition, comparator construction, index dates, outcome definitions, confounder adjustment, missing-data handling and sensitivity analyses.

### 2.8 Qualitative and yarning analysis (Objectives 1 & 2)

We will employ a critical realist approach aligned with Indigenous methodologies at a knowledge interface ^12, 18, 19^. Analysis will proceed iteratively through: (i) descriptive coding of reported changes in outcomes or implementation; (ii) analytical resolution of data into key components (e.g., actors, actions, contexts); (iii) abduction, i.e. redescribing grouped outcomes/components using substantive Indigenous and non-Indigenous theories; (iv) retroduction to identify implicit/explicit structures and mechanisms for impact, sustainability and scale; and (v) abstraction/comparison to synthesise program theories across interview transcripts ^20^.

### 2.9 Economic evaluation (Objective 3)

We will estimate the following:

- **Return-on-investment (ROI):** comparing intervention costs with hospitalisation cost offsets for HHAN versus comparators. If no positive cost offset is observed, ROI will not be reported.
- **Cost-effectiveness analysis (CEA):** estimating incremental cost-effectiveness ratios (ICERs) such as cost per emergency department presentation or hospital admission prevented. Bootstrapping will be used to improve sample size where needed; results will be presented on the cost-effectiveness plane and accompanied by sensitivity analyses.
- **Exploratory perspectives:** broader government and societal perspectives of cost and outcomes will be explored subject to data availability.

### 2.10. Multi-methods integration

Quantitative and qualitative strands will first be analysed and reported independently, then examined side-by-side to identify convergences/divergences in patterns of impact and implementation. To assess the extent to which our findings are people-centred and culturally grounded, we will interpret integrated findings through Indigenous paradigms of health and wellbeing and the WHO integrated, people-centred health services (IPCHS) framework ^**11**, **21**^.

### 2.11 Indigenous data governance, sovereignty, and cultural safety

Analyses of data relating to Aboriginal and Torres Strait Islander participants will be led by Aboriginal and Torres Strait Islander investigators and supported by an Aboriginal Advisory Committee. This governance will guide analysis plans, interpretation, and dissemination, ensuring culturally safe processes and respect for data sovereignty.

### 2.12 Data management and security

Linked HSDS data will be accessed and analysed within the Advanced Secured Analytics Lab at the New South Wales’ Government’s Data Analytics Centre. Remaining data will be stored securely at the Sydney Local Health District and The George Institute for Global Health, consistent with approved protocols. Participant lists will be stored in password-protected files; only HREC-approved personnel will have access. Study files will be retained for at least five years per research conduct codes ^**22**^.

### 2.13 Ethics and dissemination

Ethics approval was granted by the Aboriginal Health and Medical Research Council of NSW HREC (2137/23) and the Sydney Local Health District HREC (X23-0452 and 2023/ETH02445). Approval for data linkage, including waiver of consent for HSDS analyses, was granted by the NSW Population & Health Services Research Ethics Committee (2024/ETH01094). Findings will be disseminated through peer-reviewed publications, conference presentations, policy and service briefings, partner forums, community-facing outputs, and Aboriginal governance and advisory channels before or alongside broader dissemination where appropriate.

### 2.14 Team positionality and governance

The team’s diverse disciplinary and cultural expertise (including Aboriginal and Torres Strait Islander leadership) support co-interpretation and culturally safe evaluation. We have provided an overview of how we will use both Indigenous and Western analytical methods at a knowledge interface in Appendix 3.

## 3. Discussion

This protocol addresses a key concern for health managers and researchers: how to evaluate a complex, equity-oriented integrated care program without reducing it to a narrow before-and-after outcomes study. HHAN operates across networks of relationships involving individuals, organisations and institutions. Interactions across these networks are often non-linear and present emergent phenomena that demand a methodological approach capable of explaining how and why outcomes arise under specific contextual configurations ^23, 24^. The protocol therefore stands out by combining a planned counterfactual observational analysis with implementation inquiry, yarning, critical realist analysis, Indigenous data governance and economic evaluation before results are known.

By pairing counterfactual quantitative methods with qualitative study designs and yarning, the evaluation foregrounds relationality, cultural safety and explanatory inference. This is valuable because commissioners need more than evidence that service utilisation changed; they need to understand which mechanisms are activated, which contexts support or dampen them, and whether the model can be sustained or scaled without diluting culturally grounded practice.

### 3.1 Anticipated limitations and mitigations

First, as a non-randomised evaluation operating in open systems, residual confounding is likely despite weighting and double-adjustment. We will address this through sensitivity analyses and by using critical realist theory-building to identify unmeasured contextual conditions and reasoning processes that plausibly generate observed patterns (supporting retroductive inference). Second, data completeness and linkage constraints may limit statistical power and precision; these will be reported transparently with implications for inference. Third, cultural safety and data sovereignty will be maintained through governance by Aboriginal and Torres Strait Islander investigators and an Aboriginal Advisory Committee, and through co-interpretation with team members from multiple cultural heritages.

### 3.2 Implications

The study will provide decision-makers with: (i) evidence on who benefits and in which contexts; (ii) an account of how mechanisms drive impact; (iii) a value-for-money assessment relevant to budgeting, commissioning and scale-up; and (iv) a transparent example of integrating Indigenous and Western knowledge systems in applied health-service evaluation. Together, these deliverables can inform how HHAN and similar initiatives optimise service integration, sustain gains and expand without diluting cultural integrity or people-centredness.

## Ethics and dissemination

Ethics approval has been granted by the Aboriginal Health and Medical Research Council of NSW HREC (2137/23), Sydney Local Health District HREC (X23-0452 and 2023/ETH02445), and the NSW Population and Health Services Research Ethics Committee for data linkage, including waiver of consent for linked administrative data (2024/ETH01094). Findings will be shared through peer-reviewed publications, policy and service briefings, partner forums, community-facing outputs and Aboriginal governance channels.

## Trial registration number

Not applicable; this is an evaluation protocol and not a clinical trial.

## Author Approval

All authors have read and approved this protocol

## Data Availability Statement

Not applicable as this is a study protocol

## Declarations

### Consent for publication

Not applicable.

### Data availability

Linked administrative data (HSDS) are held in the NSW Government secure access environment and are not publicly available due to legal and privacy restrictions. De-identified analytic codebooks and analysis scripts for program data will be available upon reasonable request to the corresponding author and subject to approvals by the data custodians.

### Authors’ contributions (CRediT taxonomy)

- **Conceptualisation:** KY, FM, AC, MC, ERA, JC, KH, KBB, TM, MM, AT, JL, IT, SF, GS, SJ, CHS, PF, SW, RL, BG, LH, EM, JE, HL
- **Methodology and formal analysis (planned):** ERA, MC (economics), AC (statistics), KY, FM, JE (critical realist analysis), TM, MM, KH, KY, JC, KBB, HL (Indigenous methods and knowledge interface)
- **Writing** – original draft: KY
- **Writing** – review & editing: All authors
- **Project evaluation oversight:** HL

## Reporting guidelines

This manuscript has been prepared as a study protocol in accordance with BMJ guidance for protocol submissions. Because the planned evaluation includes observational analyses using routinely collected linked data, qualitative and yarning methods, and an economic evaluation, the subsequent reporting of completed findings will be guided by relevant EQUATOR resources, including STROBE and RECORD for observational/routine-data components and CHEERS for the economic evaluation component. These completed-study checklists are not presented as protocol checklists.

## Patient and public involvement

Patient, carer, provider, community and Aboriginal governance input is built into the design through yarning, qualitative interviews, partner engagement, Aboriginal and Torres Strait Islander investigator leadership and an Aboriginal Advisory Committee.

## Protocol registration

Not applicable; this evaluation is not a clinical trial.

## Acknowledgements

Not applicable.

## References

1. Eastwood J, Kemp L and Jalaludin B. “Being Alone and Expectations Lost”: A Realist Theory of Neighborhood Context, Stress, Depression, and the Developmental Origins of Health and Disease. SAGE Open 2018; 8. DOI: 10.1177/2158244018763004.

2. Subash T, Peter G, Nancy R, et al. Are adverse childhood experiences (ACEs) the root cause of the Aboriginal health gap in Australia? BMJ Global Health 2024; 9: e014901. DOI: 10.1136/bmjgh-2023-014901.

3. Eastwood JG, De Souza DE, Shaw M, et al. Designing Initiatives for Vulnerable Families: From Theory to Design in Sydney, Australia. Int J Integr Care 2019; 19: 9. 20190725. DOI: 10.5334/ijic.3963.

4. Todd K, Eastwood JG, Fotheringham P, et al. Using Geospatial Analysis to Inform Development of a Place-Based Integrated Care Initiative: The Healthy Homes and Neighbourhoods Experience. Int J Integr Care 2021; 21: 23. 20210621. DOI: 10.5334/ijic.5430.

5. Baum S and Gleeson B. Space and Place: Social Exclusion in Australia’s Suburban Heartlands. Urban Policy and Research 2010; 28: 135–159. DOI: 10.1080/08111141003693133.

6. Eastwood JG, Mukumbang FC, De Souza D, et al. Formulating Initial Program Theories of the Healthy Homes and Neighbourhoods Integrated Care Initiative. Int J Integr Care 2022; 22: 12. 20221121. DOI: 10.5334/ijic.6421.

7. Eastwood J, Barmaky S, Hansen S, et al. Refining Program Theory for a Place-Based Integrated Care Initiative in Sydney, Australia. Int J Integr Care 2020; 20: 13. 20200928. DOI: 10.5334/ijic.5422.

8. Health NMo. Creating Healthy Homes and Neighbourhoods, https://www.health.nsw.gov.au/innovation/2019awards/Pages/hhan.aspx (accessed 9 June 2020).

9. Barmaky S HS, Miller E, Tennant E, Ratcliff S, Eastwood J. A realist evaluation of Healthy Homes and Neighbourhoods’ place-based initiative in an inner city public housing estate in Sydney International Journal of Intergrated Care 2018.

10. Eastwood J. Big Data Evaluation of an Integrated Care Initiative for Vulnerable Families. International Journal of Integrated Care 2019; 19. DOI: 10.5334/ijic.s3084.

11. Bartlett C, Marshall M and Marshall A. Two-Eyed Seeing and other lessons learned within a co-learning journey of bringing together indigenous and mainstream knowledges and ways of knowing. Journal of environmental studies and sciences 2012; 2: 331–340. DOI: 10.1007/s13412-012-0086-8.

12. Nancy Clark, Janice Duddy, Madeline Gallard, et al. Using a Two-Eyed Seeing Approach to Realist Evaluation in a Community-Based Research Project, https://paninbc.ca/wp-content/uploads/2020/06/Making-it-Work-CAHR-Poster-2020.pdf (2020).

13. Department of Communities and Justice (NSW). Data Contained in the Human Services Dataset (HSDS).

14. Malterud K, Siersma VD and Guassora AD. Sample Size in Qualitative Interview Studies: Guided by Information Power. Qual Health Res 2016; 26: 1753–1760. 20160710. DOI: 10.1177/1049732315617444

15. StataCorp. Stata Statistical Software: Release 18. College Station, TX: StataCorp LLC., 2023.

16. SAS Institute Inc. SAS/STAT® 9.4: User’s Guide. Cary, NC: SAS Institute Inc., 2023.

17. R Core Team. R: A Language and Environment for Statistical Computing. Vienna, Austria: R Foundation for Statistical Computing; 2025. Available from: https://www.R-project.org/

18. Nakata M. The Cultural Interface. The Australian Journal of Indigenous Education 2007; 36: 7–14. DOI: 10.1017/S1326011100004646.

19. Bartlett C, Marshall M and Marshall A. Two-Eyed Seeing and other lessons learned within a co-learning journey of bringing together indigenous and mainstream knowledges and ways of knowing. Journal of environmental studies and sciences 2012; 2: 331–340. DOI: 10.1007/s13412-012-0086-8.

20. Danermark B, Ekström M and Karlsson JC. Explaining Society: Critical Realism in the Social Sciences Routledge, 2019.

21. World Health Organization. Framework on integrated, people-centred health services.

22. Australian Research Council. Codes and Guidelines https://www.arc.gov.au/about-arc/program-policies/research-integrity/codes-and-guidelines#Responsible (2018).

23. Amagoh F. Systems and Complexity Theories of Organizations. Global Encyclopedia of Public Administration, Public Policy, and Governance. 2016, pp.1–7.

24. Condorelli R. Complex Systems Theory: Some Considerations for Sociology. Open Journal of Applied Sciences 2016; 06: 422–448. DOI: 10.4236/ojapps.2016.67044.

